# Prevalence, awareness and factors associated with hypertension among adults in rural south-western Uganda: a cross-sectional study

**DOI:** 10.1101/2024.09.03.24313036

**Authors:** Grace Kansiime, Edwin Nuwagira, Paul Stephen Obwoya, Joseph Baruch Baluku, Michael Kanyesigye, Christine Twesiime, Peter Ssebutinde, Rose Muhindo, Anthony Muyingo, Keneth Kananura, Pliers Denis Tusingwire, Esther C. Atukunda, Robert Kalyesubula, Francis Bajunirwe, Anthony Muiru

**Affiliations:** Department of Medicine, Mbarara University of Science and Technology, Mbarara, Uganda; Division of Pulmonology, Kiruddu National Referral Hospital, Kampala, Uganda; Global Health Collaborative, Mbarara University of Science and Technology, Mbarara, Uganda; Ministry of Health, Mbarara District Local Government; Department of Medicine, Mbarara Regional Referral Hospital, Mbarara, Uganda; School of Medicine, Kabale University, Kabale, Uganda; Department of Pharmacy, Mbarara University of Science and Technology, Mbarara, Uganda; Department of Physiology and Department of Medicine, Makerere University College of Health Sciences, Kampala, Uganda; Department of Community Health, Mbarara University of Science and Technology, Mbarara, Uganda; School of Medicine, University of California, San Francisco

**Author notes:** Equal contribution (Francis Bajunirwe, Anthony Muiru). **Corresponding author:** Grace Kansiime, Department of Medicine, Mbarara University of Science and Technology, PO Box 1410, Mbarara, Uganda.

**Keywords:** Blood pressure, Cardiovascular disease, Hypertension, Rural Uganda, Sub-Saharan Africa

## Abstract

**Background:** Hypertension is the leading cause of preventable deaths globally, yet there have been inconsistent reports on its burden and risk factors in rural Sub-Saharan Africa. This study aimed to assess the prevalence, awareness, and risk factors associated with hypertension among adults in a rural community in southwestern Uganda.

**Methods:** A baseline survey was conducted as part of an ongoing implementation science cohort study in Ngango, a rural parish in the Mbarara district of southwestern Uganda. The study included adults aged 18-79 years from eleven villages. Research assistants and community health workers visited homes to enroll consenting adults. Data collection involved administering the WHO STEPS questionnaire, which gathered demographic information, behavioral characteristics, and lifestyle data, including tobacco and alcohol use, salt intake, fruit and vegetable consumption, and physical activity. Participants were also asked about prior blood pressure (BP) measurements. BP readings were taken three times, two minutes apart, along with anthropometric measurements. Hypertension was defined as BP ≥140/90 mmHg, based on the average of the last two readings, or self-reported use of antihypertensive medication. The primary outcome was the prevalence and factors associated with hypertension, assessed using logistic regression. Secondary outcomes included hypertension awareness and the proportion of participants with controlled hypertension.

**Results:** A total of 953 adults were enrolled. The median age was 43 years, with most participants being female (61.5%). Hypertension prevalence was 27.3% (260/953). Among those with hypertension, 61.5% were unaware, 27.7% were on treatment, and 65.3% had controlled BP. Despite 66.8% of participants reporting physical activity, 63.7% were overweight. Factors associated with hypertension were age > 40 years (OR 2.26, 95% CI 1.53-3.33; p <0.001), consuming <3 servings of fruit or vegetables per week (OR 1.62, 95% CI 1.11-2.35, p = 0.012), overweight (OR 1.57, 95% CI 1.05-2.34, p = 0.028), and obesity (OR 2.73, 95% CI 1.80-4.15, p <0.001).

**Conclusion:** The prevalence of hypertension in rural southwestern Uganda is high, despite a relatively young and physically active population, indicating the need for targeted interventions.

**Key Points:**

- More than 1 in 4 adults in rural Uganda have hypertension, despite high levels of physical activity (67%)
- Hypertension awareness and control rates remain below the global targets
- Our data shows that interventions targeting modifiable risk factors are urgently needed to reduce rural hypertension burden.

## Introduction

Hypertension is the leading cause of preventable deaths worldwide.(1) According to the World Health Organization (WHO), the prevalence of hypertension is highest in sub-Saharan Africa, and the average blood pressures in this region are much higher than global averages.(2) An estimated 10.8 million deaths annually are associated with high blood pressure and of these, 88% occur in low- and middle-income countries (LMICs).(3) However, it is estimated that most people with hypertension in rural sub-Saharan Africa are unaware and untreated while those who are diagnosed are not well controlled.(4, 5)

In Uganda, the prevalence of hypertension varies greatly by geographic region (6) with a disproportionately increasing burden reported in rural areas yet most studies on hypertension in Uganda have been conducted in urban areas(7, 8) or hospital settings.(9, 10) Population-based studies in rural areas are needed. To fill the knowledge gap on the epidemiology of hypertension in rural Uganda, we conducted a cross-sectional study, assessing the prevalence, awareness, and factors associated with hypertension among adults living in a rural community in south-western Uganda.

## Methods

### Study Design and setting

This cross-sectional survey is part of an ongoing mixed methods implementation science cohort study known as Comprehensive Hypertension Improvement in Rural Uganda (CHIRU study). The CHIRU study aims to integrate community health workers (CHWs) also called village health teams (VHTs) in hypertension management in Ngango parish, in Kagongi sub-county, Western Uganda. A parish is an administrative unit in Uganda consisting of multiple villages. We included adult residents from eleven villages of Ngango Parish between September 25, 2023 and November 30, 2023. Ngango Parish is one of the six parishes in Kagongi sub-county in Mbarara district (Figure 1) which is approximately 270km southwest of Kampala, Uganda’s capital city. This Parish was purposively selected because it houses the only public health facility whose health workers were part of the original cohort study. The community dwellers are mostly subsistence farmers.

**Figure 1:**
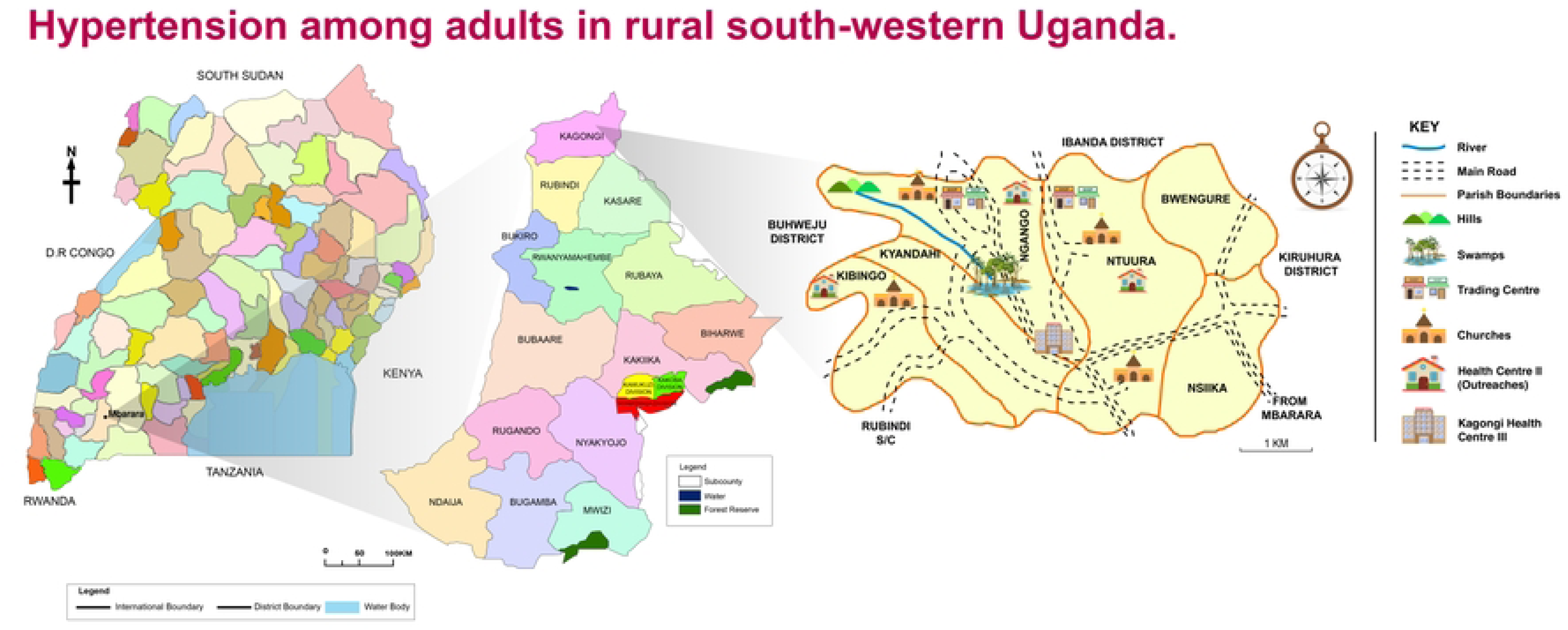
**Map of Mbarara district showing Kagongi sub-county and Ngango parish.**

### Community Engagement and Study Population

Ngango Parish consists of 11 villages, each supported by two community health workers (CHWs) who were introduced to the research team by their coordinator at the local health center. These CHWs are community members trained by the government to link the community with health facilities. They play a vital role in health education, providing first aid, and managing common health issues such as diarrhea, malaria, and pneumonia in children, (11, 12), and HIV in adults(13). Before the study began, the CHWs shared population records with the research team, showing that Ngango Parish has a total population of 4,355 people, of which 2,133 (49%) are adults aged 18 years and older, including 991 males and 1,142 females.

The study targeted adults aged 18-79 years who had lived in Ngango Parish for at least six months and provided informed consent. Before data collection, the research team, including the principal investigator (PI) and research assistants, held meetings with local council leaders and CHWs to inform them about the study and to schedule visits to each village. Data collection was planned for afternoons from Monday to Saturday to avoid disrupting the community’s usual farming and church activities.

To maximize participation, local council leaders and CHWs used local radios (megaphones) and announcements at communal gatherings, such as places of worship, to inform residents. The research team conducted house-to-house visits across all 11 villages (including trading centres/shops), recruiting all eligible adults found at home for the study.

### Study procedures

#### Training and Data Collection

Community health workers (CHWs) received one week of training on hypertension management using the CDC’s hypertension management curriculum. (14) Research assistants underwent a 3-day training focused on study procedures and the administration of the STEPS tool. For data collection, each trained research assistant was paired with a CHW to facilitate the process, with data being collected from participants in their homes.

#### Study measurements

Data were collected using the WHO STEPS tool for non-communicable diseases (NCDs) surveillance during household visits.(15) In this stepwise approach to chronic disease risk factor surveillance, an interviewer-administered questionnaire was used, consisting of three steps:

**STEP 1:** We gathered data on demographic, socio-economic, and behavioral characteristics, including lifestyle factors such as tobacco use, alcohol consumption, salt intake, fruit and vegetable consumption, and physical activity. Current alcohol users were defined as participants who had consumed alcohol in the past 30 days. Salt intake was evaluated by asking how often participants added salt to their food before tasting or during meals and classified as ‘Too much’ if they added salt at table always, ‘Enough’ if they rarely added salt at table and as ‘Little’ if they never added salt at table. Physical activity was assessed by inquiring about daily work, typical modes of transportation, and time spent sitting each day. Additionally, we asked whether participants had ever had their blood pressure measured.

**STEP 2:** Physical measurements were conducted, including weight, height, waist and hip circumferences, and blood pressure. Blood pressure was measured on the left arm while the participant was seated with legs uncrossed, back supported, after at least 5 minutes of rest using an appropriately sized cuff and a battery-powered digital blood pressure machine (Omron 3 Series, Omron, Kyoto, Japan). The cuff (size 22-42cm) was applied to the arm, and the machine automatically took three BP measurements, two minutes apart; the average of the last two readings was recorded.

Height was measured manually with the participant standing upright against a wall, using a tape measure to derive the height. Measurements were taken with the participant barefoot, standing with their back and head against the wall, heels together, and stretched to the fullest. Height was recorded to the nearest centimeter. Weight was measured using a pre-calibrated digital scale (SECA 877), with the participant in light clothing and barefoot, and recorded to the nearest tenth of a kilogram.

**STEP 3:** Blood sugar levels were measured using On Call®Plus glucometers (ACON USA), which were calibrated each morning before data collection. Participants with a random blood sugar (RBS) reading >11.1mmol/L were scheduled for a follow-up test the next morning after an overnight fast.

We used Research Electronic Data Capture (REDCap, Vanderbilt University), a cloud-based secure HIPAA-compliant system, for data collection and management.(16)

#### Covariates

Descriptive analyses included baseline demographics such as age, sex, education level, marital status, and occupation. Comorbidities, including diabetes mellitus (DM), were considered, along with body mass index (BMI), dietary salt intake, fruit and vegetable consumption. Lifestyle factors and social habits like physical activity, smoking, and alcohol consumption were also included.

#### Study Outcomes

The primary outcome of the study was the prevalence of hypertension among adults in the rural community. Hypertension was defined as a blood pressure ≥140/90mmHg per the 2020 International Society of Hypertension (ISH) Global Hypertension Practice Guidelines(17) or taking anti-hypertension medication. Using the same guidelines, blood pressure readings were grouped as normal blood pressure for SBP<130 mmHg and DBP <85 mmHg, high normal blood pressure (Prehypertension) for SBP 130–139 and/or DBP 85–89, grade 1 hypertension SBP 140– 159 mmHg and/or DBP 90–99 mmHg, grade 2 hypertension SBP ≥160 mmHg and/or DBP ≥100 mmHg. All definitions were based on an average of the last two of the three blood pressure readings taken for each participant. Secondary outcomes included the awareness of hypertension status, the proportion of participants with controlled hypertension, and the identification of factors associated with hypertension. Hypertension awareness was defined as the proportion of participants who were aware that they had raised blood pressure among those found to have hypertension in the study while controlled hypertension was defined as BP <140/90mmHg among those who were taking anti-hypertension medications. Diabetes mellitus (DM) was defined as a fasting blood sugar ≥ 7.0mmol/l and/or reported use of antidiabetic medications. High BMI was defined as a BMI ≥ 25 kilograms/meters^2^

#### Data analysis

Descriptive statistics were used to summarize the data: means and standard deviations for normally distributed continuous variables, and medians with interquartile ranges for skewed continuous variables. Categorical variables were reported as counts and percentages. Baseline characteristics were compared between participants with and without hypertension using the Wilcoxon rank-sum test for continuous variables and the chi-square (χ²) test for categorical variables. Hypertension prevalence was calculated as the percentage of participants with a blood pressure ≥140/90 mmHg or those taking prescribed antihypertensive medications.

For identifying factors associated with hypertension, we selected variables based on domain knowledge, including age, sex, body mass index, physical activity, education level, occupation, comorbidities like diabetes mellitus, salt and fruit intake, smoking, and alcohol use. Binary logistic regression was performed at both univariate and multivariable levels. Variables with a p-value <0.23 in univariate analysis and those considered biologically plausible including diabetes mellitus, physical inactivity, diet (fruit/vegetable and salt consumption) were included in the multivariable analysis. Results are presented as crude and adjusted odds ratios with 95% confidence intervals and p-values. Factors with p<0.05 were considered statistically significant.

We used the Strengthening The Reporting of Observational Studies in Epidemiology (STROBE) cross-sectional study checklist when writing our report(18).

#### Ethical considerations

The protocol received ethical approval from the Mbarara University of Science and Technology research and ethics committee (MUST-2023-382) and the Uganda National Council for Science and Technology (HS2357ES). All study participants gave written informed consent to participate in the study and the study was conducted per the Declaration of Helsinki 2013.

## Results

Between September and November 2023, we screened 972 adults in Ngango Parish for eligibility and included 953 participants for this study as shown below (Figure 2).

**Figure 2:**
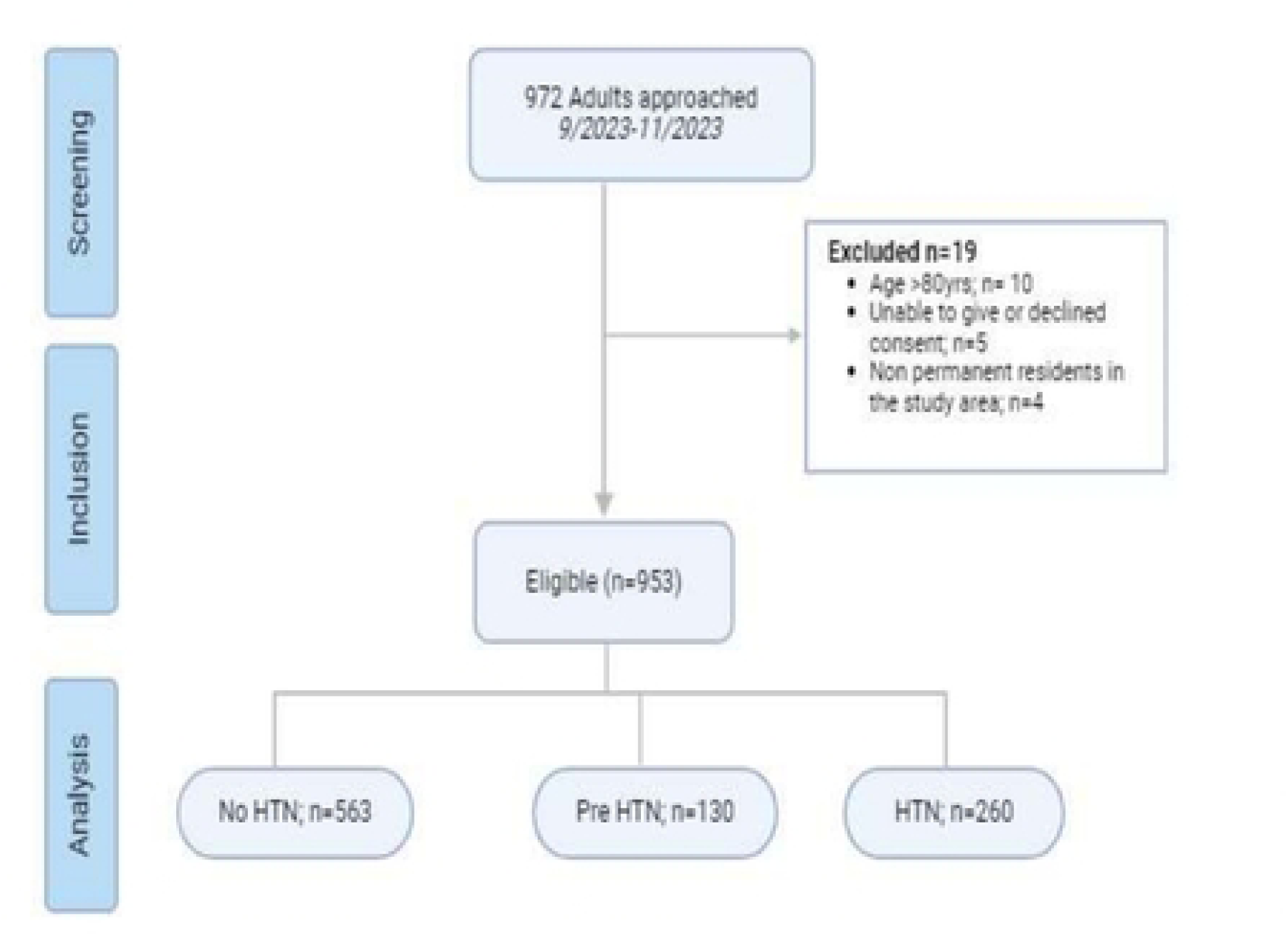
**Study flow diagram**

### Sociodemographic Characteristics of Participants

Of the 953 participants, 586 (61.5%) were female, and the median (IQR) age was 42.6 (30.1-56.7) years. Six out of every 10 individuals were overweight (BMI >24.9 kg/m^2^) although 66.8% reported being physically active. Less than half of the participants 415/953 (43.5%) recalled having ever had their BP measured. The median (IQR) random blood sugar of the participants was 5.7 (4.9-6.7) with only 30 (3.1%) participants having diabetes mellitus. Other baseline characteristics are shown on Table 1. Participants were evenly distributed among all participating villages (Figure s1)

**Table 1:** Baseline characteristics of study participants.

|  |  |  |  |  |
| --- | --- | --- | --- | --- |
| Physical activity n (%) |  |  |  |  |
| No | 316 (33.2) | 231 (33.3) | 85 (32.7) |  |
| Yes | 637 (66.8) | 462 (66.7) | 175 (67.3) |  |
|  |  |  |  | 0.85 |
| Current Alcohol Use n (%) |  |  |  |  |
|  | 171 (17.9) | 121 (17.5) | 50 (19.2) |  |
|  | 782 (82.1%) | 572 (82.5%) | 210 (80.8%) |  |
| Yes |  |  |  | 0.53 |
| No |  |  |  |  |
| Current smoker n (%) |  |  |  |  |
| No | 863 (90.6%) | 633 (91.3%) | 230 (88.5%) | 0.18 |
| Yes | 90 ( 9.4%) | 60 ( 8.7%) | 30 (11.5%) |  |
|  | Total<br>N=953 | No HTN<br>N=693 | HTN<br>N=260 | p-value |
| **Sex n (%)** |  |  |  |  |
| Male | 367 (38.5) | 270 (39.0) | 97 (37.3) | 0.64 |
| Female | 586 (61.5) | 423 (61.0) | 163 (62.7) |  |
| **Age median (IQR)** | 43 (30-57) | 39 (28-54) | 52 (35-65) | <0.001 |
| **Marital status N= 921 n (%)** |  |  |  |  |
| Single | 136 (14.8%) | 109 (16.3) | 27 (10.6) | 0.026 |
| Married | 655 (71.1%) | 474 (71.1) | 181 (71.3) |  |
| Separated/Widowed | 130 (14.1%) | 84 (12.6) | 46 (18.1) |  |
| **Own a mobile phone n (%)** |  |  |  |  |
| No | 444 (46.6) | 325 (46.9) | 119 (45.8) | 0.76 |
| Yes | 509 (53.4) | 368 (53.1) | 141 (54.2) |  |
| **Monthly income, median (IQR), USD** | 39.5 (15.8-79.0) | 39.5 (18.4-65.8) | 39.5 (15.8-79.0) | 0.26 |
| **Education level N=946 n (%)** |  |  |  |  |
| No Formal Education | 158 (16.7) | 98 (14.2) | 60 (23.3) | <0.001 |
| Primary | 555 (58.7) | 404 (58.6) | 151 (58.8) |  |
| Secondary and Above | 233 (24.6) | 187 (27.1) | 46 (17.9) |  |
| **BMI median (IQR) kg/m<sup>2</sup>** | 26.7 (23.7-30.3) | 26.3 (23.4-29.7) | 27.8 (24.5-31.6) | <0.001 |
| **BMI category\* n (%)** |  |  |  |  |
| Underweight | 28 (2.9) | 21 (3.0) | 7 (2.7) | <0.001 |
| Normal | 318 (33.4) | 249 (35.9) | 69 (26.5) |  |
| Overweight | 348 (36.5) | 260 (37.5) | 88 (33.8) |  |
| Obesity | 259 (27.2) | 163 (23.5) | 96 (36.9) |  |
| **RBS median (IQR) mmol/l** | 5.7 (4.9-6.7) | 5.7 (4.9-6.7) | 5.7 (4.8-6.8) | 0.67 |
| **Diabetes Mellitus n (%)** |  |  |  |  |
| Yes | 30 (3.1) | 16 (2.3) | 14 (5.4%) | 0.015 |
| No | 923 (96.9) | 677 (97.7) | 246 (94.6%) |  |
| **Ever had a BP measured n (%)** |  |  |  |  |
| Yes | 415 (43.5) | 265 (38.2) | 150 (57.7) | <0.001 |
| No | 538 (56.5) | 428 (61.8) | 110 (42.3) |  |
| **Reported salt intake, N=898, n (%)** |  |  |  |  |
| Too much | 90 (10.0) | 65 (9.9) | 25 (10.3) | 0.10 |
| Enough | 498 (55.5) | 379 (57.8) | 119 (49.2) |  |
| Little | 310 (34.5) | 212 (32.3) | 98 (40.5) |  |
| **Daily Fruit and Veg Servings, N=933, n (%)** |  |  |  |  |
| 1 | 142 (15.2) | 98 (14.4) | 44 (17.3) | 0.24 |
| 2 | 527 (56.5) | 378 (55.7) | 149 (58.7) |  |
| 3 | 125 (13.4) | 99 (14.6) | 26 (10.2) |  |
| ≥4 | 139 (14.9) | 104 (15.3) | 35 (13.8) |  |
| **Physical activity n (%)** |  |  |  |  |
| No | 316 (33.2) | 231 (33.3) | 85 (32.7) | 0.85 |
| Yes | 637 (66.8) | 462 (66.7) | 175 (67.3) |  |
| **Current Alcohol Use n (%)** |  |  |  |  |
| Yes | 171 (17.9) | 121 (17.5) | 50 (19.2) | 0.53 |
| No | 782 (82.1%) | 572 (82.5%) | 210 (80.8%) |  |
| **Current smoker n (%)** |  |  |  |  |
| No | 863 (90.6%) | 633 (91.3%) | 230 (88.5%) | 0.18 |
| Yes | 90 ( 9.4%) | 60 ( 8.7%) | 30 (11.5%) |  |

### Prevalence and distribution of Hypertension and pre-hypertension among study participants

The overall prevalence of hypertension in this rural community population was 27.3% (260/953) while 13.6% (130 /953) had pre-hypertension. Hypertension prevalence was similar among males and females (26.4% vs 27.8% p-value 0.64. Most of the participants had stage 1 hypertension (47%), followed by pre-hypertension (38%) and grade 2 hypertension (15%) (Figure s2).

#### Awareness and control of Hypertension

Of the 260 participants with hypertension, 160 (61.5%) were unaware that they had hypertension, 72 (27.7%) were on treatment and 47/72 (65.3%) had their blood pressure controlled. Females were more likely to be aware and on treatment for hypertension although males had better BP control among those on treatment. (Figure 3)

**Figure 3:**
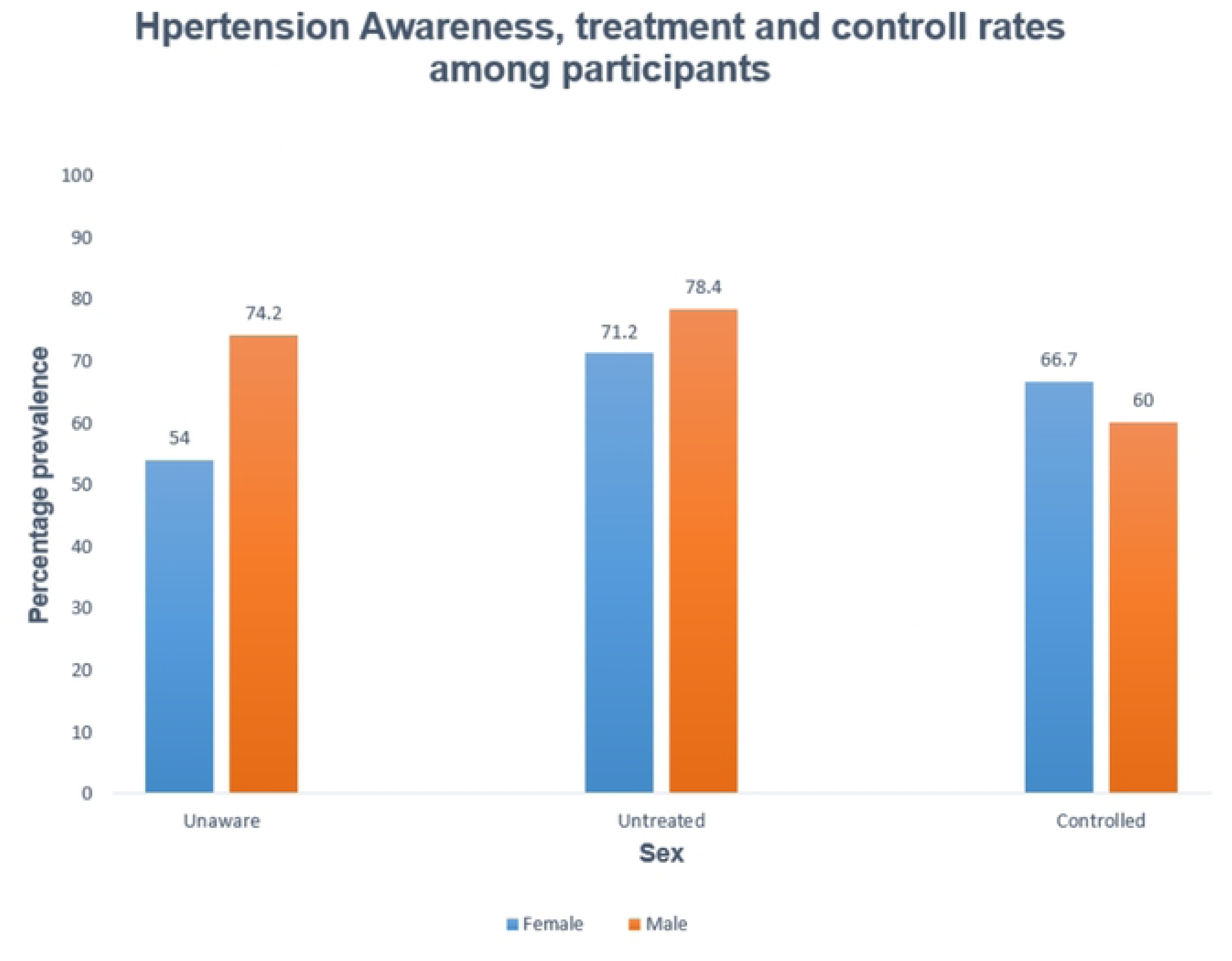
**A bar graph showing the proportion of participants aware of their hypertension status, those not on medication, and those with controlled hypertension by Sex.**

### Factors associated with hypertension among adults in Ngango Parish

Participants who were aged 40 years and above had higher odds of having hypertension compared to those below 40 (OR 2.26, 95% CI 1.53-3.33, p <0.001), consuming less than 3 fruit or vegetable servings per week (OR 1.62 95% CI 1.11-2.35, p 0.012), and higher BMI was also associated with hypertension; overweight (OR 1.57; 95% CI 1.05-2.34, p 0.028) and obesity (OR 2.73; 95% CI 1.80-4.15, p <0.001) compared to normal weight. There was no significant association between hypertension and diabetes mellitus, alcohol intake, smoking, salt /fruit / vegetable consumption or physical activity (Table 2, and figure s3)

**Table 2:**
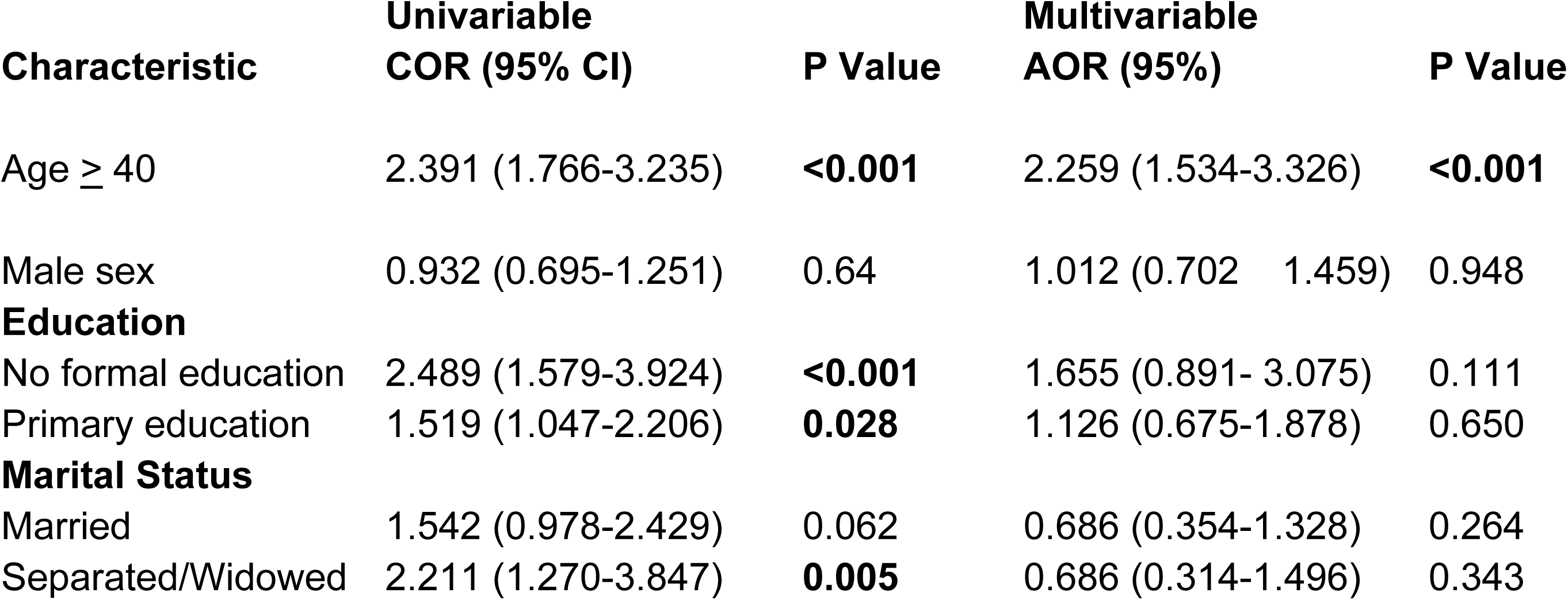
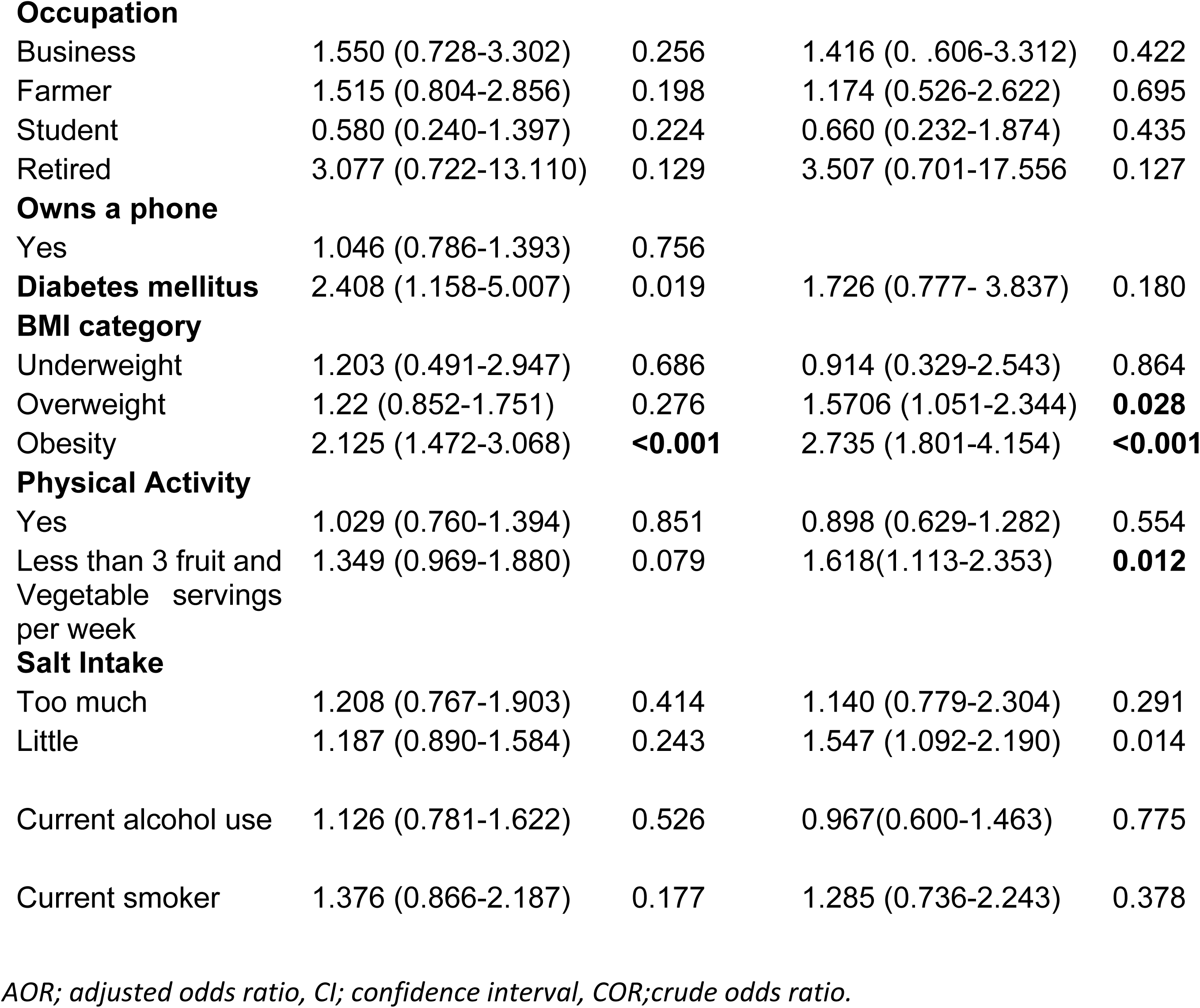
Factors associated with hypertension among adults in Ngango parish.

| Characteristic | Univariable<br>COR (95% CI) | P Value | Multivariable<br>AOR (95%) | P Value |
| --- | --- | --- | --- | --- |
| Age ≥ 40 | 2.391 (1.766-3.235) | **<0.001** | 2.259 (1.534-3.326) | **<0.001** |
| Male sex | 0.932 (0.695-1.251) | 0.64 | 1.012 (0.702 1.459) | 0.948 |
| **Education** |  |  |  |  |
| No formal education | 2.489 (1.579-3.924) | **<0.001** | 1.655 (0.891- 3.075) | 0.111 |
| Primary education | 1.519 (1.047-2.206) | **0.028** | 1.126 (0.675-1.878) | 0.650 |
| **Marital Status** |  |  |  |  |
| Married | 1.542 (0.978-2.429) | 0.062 | 0.686 (0.354-1.328) | 0.264 |
| Separated/Widowed | 2.211 (1.270-3.847) | **0.005** | 0.686 (0.314-1.496) | 0.343 |
| **Occupation** |  |  |  |  |
| Business | 1.550 (0.728-3.302) | 0.256 | 1.416 (0.606-3.312) | 0.422 |
| Farmer | 1.515 (0.804-2.856) | 0.198 | 1.174 (0.526-2.622) | 0.695 |
| Student | 0.580 (0.240-1.397) | 0.224 | 0.660 (0.232-1.874) | 0.435 |
| Retired | 3.077 (0.722-13.110) | 0.129 | 3.507 (0.701-17.556) | 0.127 |
| **Owns a phone** |  |  |  |  |
| Yes | 1.046 (0.786-1.393) | 0.756 |  |  |
| **Diabetes mellitus** | 2.408 (1.158-5.007) | 0.019 | 1.726 (0.777- 3.837) | 0.180 |
| **BMI category** |  |  |  |  |
| Underweight | 1.203 (0.491-2.947) | 0.686 | 0.914 (0.329-2.543) | 0.864 |
| Overweight | 1.22 (0.852-1.751) | 0.276 | 1.5706 (1.051-2.344) | **0.028** |
| Obesity | 2.125 (1.472-3.068) | **<0.001** | 2.735 (1.801-4.154) | **<0.001** |
| **Physical Activity** |  |  |  |  |
| Yes | 1.029 (0.760-1.394) | 0.851 | 0.898 (0.629-1.282) | 0.554 |
| Less than 3 fruit and<br>Vegetable servings<br>per week | 1.349 (0.969-1.880) | 0.079 | 1.618(1.113-2.353) | **0.012** |
| **Salt Intake** |  |  |  |  |
| Too much | 1.208 (0.767-1.903) | 0.414 | 1.140 (0.779-2.304) | 0.291 |
| Little | 1.187 (0.890-1.584) | 0.243 | 1.547 (1.092-2.190) | 0.014 |
| Current alcohol use | 1.126 (0.781-1.622) | 0.526 | 0.967(0.600-1.463) | 0.775 |
| Current smoker | 1.376 (0.866-2.187) | 0.177 | 1.285 (0.736-2.243) | 0.378 |

*AOR; adjusted odds ratio, CI; confidence interval, COR; crude odds ratio.*

## Discussion

This cross-sectional study of individuals living in rural Uganda reveals a relatively high prevalence of hypertension, a low rate of hypertension awareness, and fair hypertension control rates among those on treatment. Identified risk factors for hypertension included older age and higher BMI. While the epidemiology of hypertension has been well studied in urban populations in Uganda, there is limited data from rural areas, and thus our work fills an important knowledge gap in the field.

Although the prevalence of hypertension in our rural population was lower compared to the Ugandan national average (31.5%),(6) it is one of the highest reported in western Uganda, adding to the literature on the geographic variation in hypertension prevalence. One potential reason for these differences in prevalence is variations in BP measurement protocols across studies, which would make comparisons within studies difficult. Another more concerning reason for the higher prevalence in our study could be an increase in prevalence over time; for instance, one study from rural western Uganda found a prevalence of 14.6% in 2013 (19) compared with our findings of 27% a decade later. This increasing trend of hypertension among rural dwellers has been shown in other studies from sub-Saharan Africa.(20–22) This highlights the need to conduct longitudinal studies that include diverse communities to better understand the burden of hypertension in Uganda.

Consistent with our current understanding of hypertension pathophysiology,(23, 24) we found that older age, low fruit/vegetable consumption and higher BMI were significantly associated with hypertension in our study. We were however surprised by the high prevalence of overweight and obesity. Some have suggested that individuals living in rural Africa are physically active in their daily farming work and have not adopted the Westernized lifestyle to a greater extent.(19, 21, 25–27) We postulate that despite of the self-reported high physical activity, the high rates of overweight and obesity are driven by dietary patterns and not a sedentary lifestyle among rural dwellers. Diets in rural Uganda are characterized by monotonous high-glycemic-index staple foods and sugary foods that are associated with high BMIs.(28) In fact, 72% of participants reported having ≤2 servings of fruits and vegetables per day, far below the recommended 5 servings that have been shown to reduce all-cause mortality and CVD-related deaths.(29) Given the increasing rates of overweight and obesity among rural dwellers, more innovative ways are needed to address this key hypertension risk factor.

The observed low rates of hypertension awareness are concerning but unfortunately consistent with other studies from Africa, (30) (31) and highlights the need for ongoing strategies to address this burden hypertension. Despite the low The fact that only two out of every five individuals have ever had their blood pressure measured further underscores the extent of work needed to improve screening, treatment, and control in our settings. Despite the low rates of awareness, those who were on treatment exhibited fairly good hypertension control, as has been demonstrated in previous studies.(22) This finding highlights a crucial opportunity for improving outcomes if more individuals are made aware of their condition and receive appropriate treatment for hypertension. We leveraged an existing frame work of community health workers to rapidly screen for hypertension (visiting almost 700 households in a few months), demonstrating that this approach can be an efficient strategy to increase hypertension awareness in rural areas.

Our findings should be interpreted in the context of our study limitations. First, this is a cross-sectional study, with blood pressure measured in one encounter, which can lead to an overestimation of hypertension prevalence because we were unable to confirm the presence of elevated BP on two separate occasions. In addition, while we assessed the association between behavioral risk factors and hypertension, there is likely an element of recall bias regarding, vegetable and fruit consumption, salt intake, and physical activity. Finally, while we moved from home to home during data collection, we did convenient sampling as we only enrolled those people we found at home. However, we utilized a validated WHO data collection instrument to obtain our data and thus confident in our measurements. Moreover, the BP measurements were performed at home (cf. office measurements), minimizing the possibility of white coat hypertension as participants were likely to be more relaxed in their familiar home environments.

Despite these limitations, our study was innovative by using a home-to-home approach in the afternoons when most people are back home from their fields and included all adults found at home to get a good representation of this community. The finding that less than half of the adults in the community have had their BP measured highlights the need for accessible BP monitoring, such as community-placed machines overseen by VHTs. This would supplement the current practice of BP measurement, which is limited to health facility visits or infrequent health camps.

## Conclusion

We report a high prevalence of hypertension, a low rate of disease awareness, with a fair control among relatively young physically active individuals living in rural Uganda.

## Disclosure

The authors report no conflicts of interest in this work.

## Data Availability

The datasets used and/or analyzed during the current study are available from the corresponding author on reasonable request.

## Acknowledgment

We are profoundly grateful to Prof. Ivers Louise for her guidance and mentorship throughout the manuscript preparation. We are also immensely thankful to the research assistants and Village Health Team members for their diligent efforts and dedication. Finally, we extend our sincere appreciation to all study participants from the villages of Ngango Parish, Kagongi Sub-county, whose participation and cooperation were crucial to the success of this research.

## Author contributions

GK: Conceptualization, Data curation, Study administration, Formal analysis, Writing - original draft, Writing - review and editing; EN: Conceptualization, Writing - review and editing; PSO: Data curation, Writing - review and editing; BBJ: Conceptualization, Writing - original draft, Writing - review and editing; MK: Formal analysis, Writing - original draft, Writing - review and editing; CT: Study administration, Data curation, Writing - review and editing; PS: Data curation, Writing - review and editing; RM: Formal analysis, Writing - review and editing; AM^1^: Data curation, Methodology, Writing - review and editing; KK: Writing - original draft, Writing - review and editing; TPD: Conceptualization, Study administration, data curation, Writing - review and editing. ECA: Data curation and analysis, Writing - review and editing; RK: Conceptualization, Methodology, Writing - review and editing; FB: Conceptualization, Study administration, Writing - review and editing. AM^2^: Conceptualization, Methodology, Writing - review and editing.

